# Preventing railway suicide: learning from Australia’s success

**DOI:** 10.64898/2026.06.14.26355449

**Authors:** Phillip Cheuk Fung Law, Sangsoo Shin, Alan Woodward, Jane Pirkis, Roderick McClure, Lyndal Bugeja, Karl Andriessen, Anna Brooks, Lay San Too

## Abstract

**Objective:** To examine railway suicide trends and change points in Australia and across states/territories.

**Methods:** We identified railway suicides that occurred in Australia between 2001 and 2023, using data from the National Coronial Information System. We performed negative binomial regression analysis to examine railway suicide trends and joinpoint regression analysis to identify potential change points at both national and state/territory levels.

**Results:** Between 2001 and 2023, railway suicide in Australia declined by 54% (Incidence rate ratio [IRR] 0.98, 95% Confidence Interval [CI] 0.97 to 0.99). Substantial declines were observed in Victoria (−62%, IRR 0.97, 95% CI 0.96 to 0.99) and New South Wales (−67%, IRR 0.97, 95% CI 0.96 to 0.99), and, to a lesser extent, in Queensland (−25%, IRR 0.97, 95% CI 0.95 to 0.99). Nationally, one change point was identified. Between 2017 and 2023, the annual percent change was −8.8% (95% CI −24.5 to −3.3). In Victoria, railway suicide rates decreased annually by 13.6% between 2018 and 2023 (95% CI −38.5 to −4.3). In Western Australia, railway suicide rates increased annually by 10.3% between 2002 and 2010 (95% CI 1.5 to 58.8) and decreased annually by 5.4% between 2010 and 2023 (95% CI −19.8 to −1.8). No change point was identified for other states/territories.

**Conclusion:** Australian railway suicides have declined substantially, with this trend largely driven by reductions in Victoria and New South Wales. These findings demonstrate that railway suicide is preventable through multisectoral initiatives and suggest that interventions should be continued to reduce railway suicide.

## Introduction

Globally, suicides occurring on the railway networks represent a small proportion of all suicides, ranging from 1% to 12% (Mishara and Bardon, 2016). However, the impact of railway suicide is substantial and long-lasting, and includes traumatic experiences for family members, witnesses (e.g., train driver, railway staff, bystanders, passengers), and first responders. Railway suicides also impose substantial financial costs on railway operators (e.g., resulting from delays and cancellations of railway services) (Bardon and Mishara, 2015; Colovic, 2024).

Various interventions have been implemented to prevent railway suicide (Belur et al., 2025). Restricting access to means by installing platform screen doors/barriers at railway stations, installing fencing and removing railway crossings on open tracks, and using suicide pits has been shown to be effective in reducing railway suicides (Belur et al., 2025; Pirkis et al., 2015; Too et al., 2025), with no clear evidence of displacement to other locations (Too et al., 2025). Responsible media reporting has also been associated with a reduction in railway suicides (Etzersdorfer and Sonneck, 1998; Niederkrotenthaler & Sonneck, 2007; Sonneck et al., 1994). Railway suicides have also been shown to decrease after the installation of blue lights (Ichikawa et al., 2014; Matsubayashi et al., 2013; Matsubayashi et al., 2014). Signs at railway stations that point people to helplines may be a useful intervention (Erlangsen et al., 2023).

In Australia, multisectoral programs and partnerships have been established to reduce railway suicide. For example, a railway suicide prevention research program was implemented in Victoria between 2013 and 2016. This program was initiated in recognition of Victoria having the highest railway suicide rate in Australia and concerns raised by the public and railway sector about the traumatic impact of railway suicide. The program was developed and delivered by the University of Melbourne’s research team (see Supplementary Material Table S1 for details) in collaboration with the Victorian Department of Transport, Planning and Local Infrastructure, Victoria Police, Coroners Court of Victoria, and six railway regulators and operators. During the program period, a series of stakeholder meetings were convened to facilitate research aimed at informing railway suicide prevention strategies (Too et al., 2014; Too et al., 2015; Too et al., 2016; Too et al., 2017a; Too et al., 2017b). In 2016, the TrackSAFE Foundation identified suicide prevention as a priority for the railway industry and incorporated it into their work programs from 2017 onwards. They partnered with Lifeline Australia and developed the “Pause.Call.Be Heard” campaign to promote help seeking among railway commuters to prevent railway suicide. Between 2017 and 2018, roundtables on “Suicide Safer Railways” with relevant ministers and key stakeholders were held in three states (Victoria, New South Wales, and Queensland) to develop recommendations for preventing railway suicide. Since 2017, preventive interventions have been implemented actively across Australian railway networks (e.g., installing fencing at high-risk railway locations, “Pause.Call.Be Heard” campaign; see Supplementary Material Table S1 for the timeline and details of these initiatives/interventions). Despite the implementation of these interventions, no study has examined whether railway suicide has declined in Australia and across states/territories. This study aimed to examine trends in railway suicide rates at national and state/territory levels. It also aimed to identify change points to assess whether railway suicide rates shifted around the introduction of these initiatives/interventions.

## Methods

### Railway suicide counts

We identified suicides occurring in Australian railway environment (railway suicide) from the National Coronial Information System (NCIS). The NCIS is an online database of Australian and New Zealand coronial information (Dunstan, 2019). It contains suicide data from July 2000 for all states/territories, except Queensland where data are available from January 2001. Each record includes coded data (sociodemographic and death information) and full-text reports (the police summary of circumstances, autopsy report, toxicology report, and the coroners’ findings). We extracted data on incident year and state/territory where the railway suicide occurred. We included railway suicides that occurred between January 2001 and December 2023 and excluded cases with unknown year of incident.

### Population estimates

We obtained annual resident population estimates from 2001 to 2023 at national and state/territory levels from the Australian Bureau of Statistics publicly available source (Australian Bureau of Statistics, 2025).

### Statistical analysis

We estimated railway suicide trends between 2001 and 2023 for Australia and across states/territories using negative binomial regression model. Railway suicide counts were included as the outcome and continuous incident year as the exposure, with log population number as the offset term. Incidence rate ratio (IRR) and 95% Confidence Interval (CI) were reported. This analysis was performed using STATA/SE 15.1.

We performed joinpoint regression to identify change points in Australia and across states/territories using railway suicide rates. The models were fitted using log transformation of railway suicide rate as the outcome and incident year as the exposure. The results were therefore interpreted as the annual percent change (APC) and reported with their 95% CI. We conducted the regression analysis from a minimum of zero to maximum of five join points and selected the best fitting model based on the weighted Bayesian Information Criterion (BIC). A lower weighted BIC indicates better model fit. Data analysis was conducted using the National Cancer Institute’s Joinpoint Regression Program (version 6.0.1).

## Results

### Railway suicide trends

Railway suicide accounted for 2.7% of all suicides in Australia between 2001 and 2023, with an average annual rate of 0.31 per 100,000 persons (Table 1). Victoria had the highest railway suicide rate (5.5% of all suicides, 0.56 per 100,000 persons), followed by New South Wales (2.8%, 0.29 per 100,000 persons), and Western Australia (1.9%, 0.25 per 100,000 persons). There was a significant decline in Australian railway suicide rates, corresponding to an average 2% reduction per year (IRR 0.98, 95% CI 0.97□0.99) and a 54% reduction between 2001 and 2023. Similarly, a significant decrease in railway suicide rates was observed in Victoria (an average 3% decline per year; IRR 0.97, 95% CI 0.96□0.99; −61.7% over time), New South Wales (an average 3% decline per year; IRR 0.97, 95% CI 0.96□0.99; −66.7% over time), and Queensland (an average 3% decline per year; IRR 0.97, 95% CI 0.95□0.99; −25.2% over time). Figure 1 shows the railway suicide rates at the national and state/territory levels between 2001 and 2023.

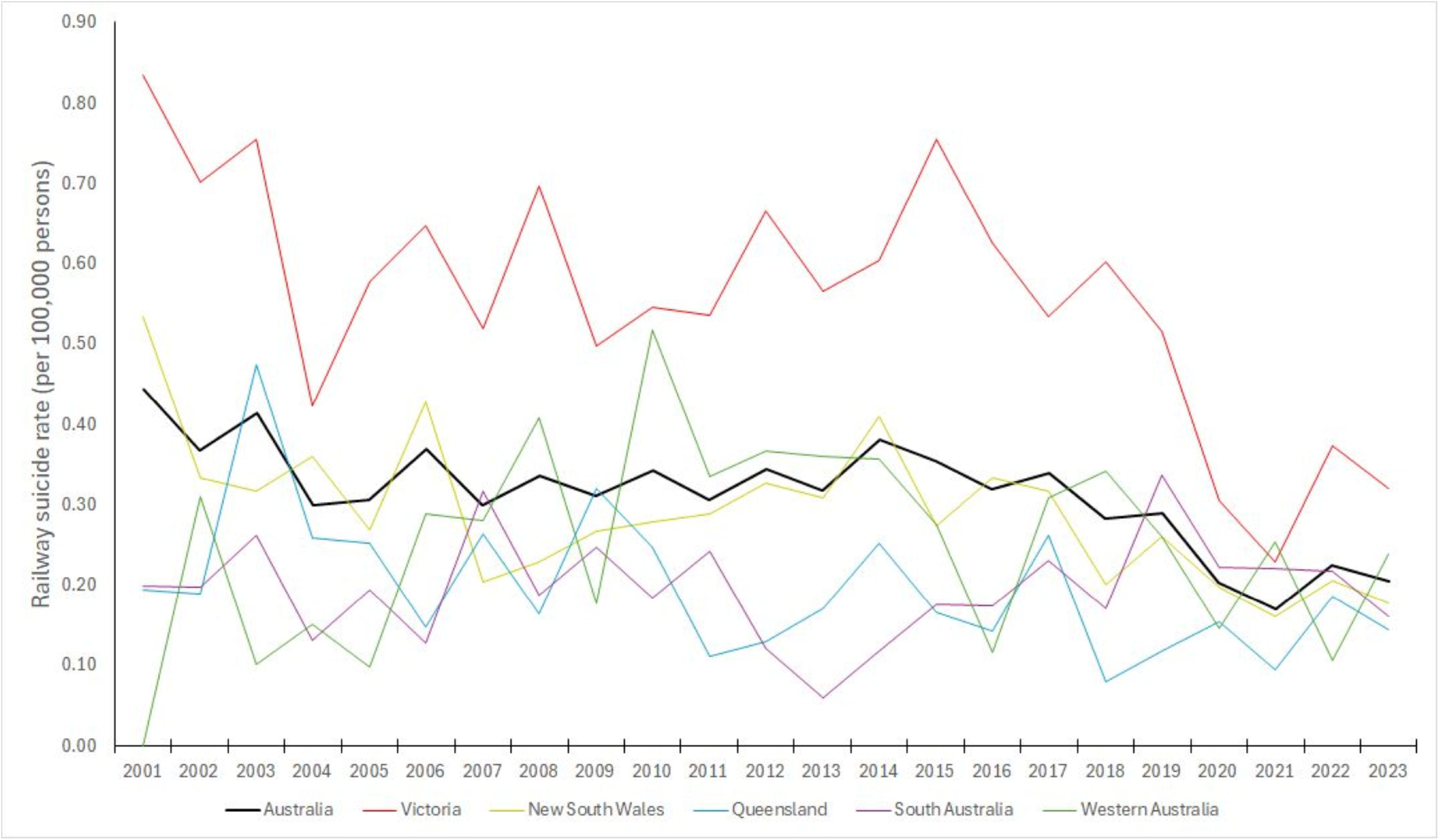

**Table 1.**
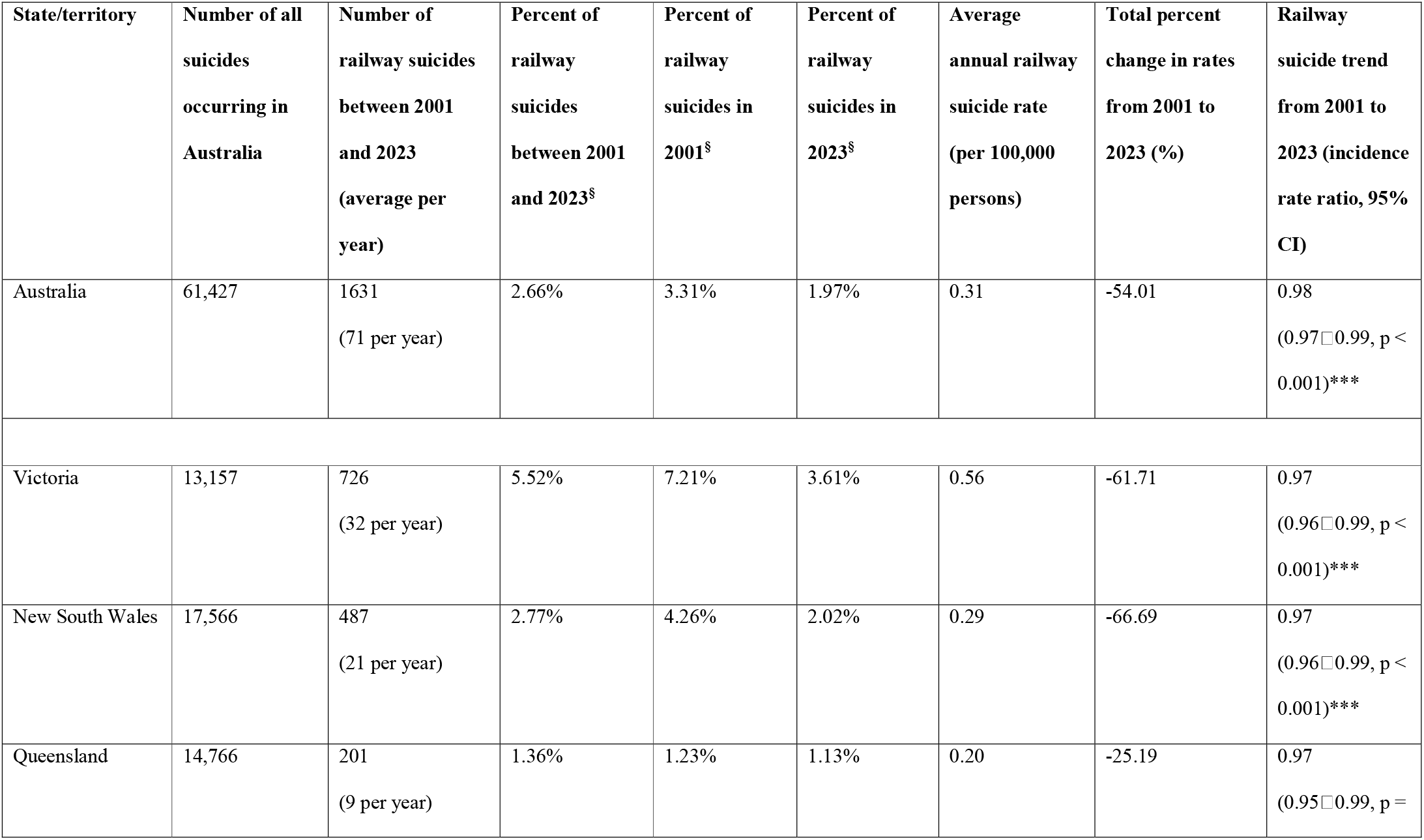
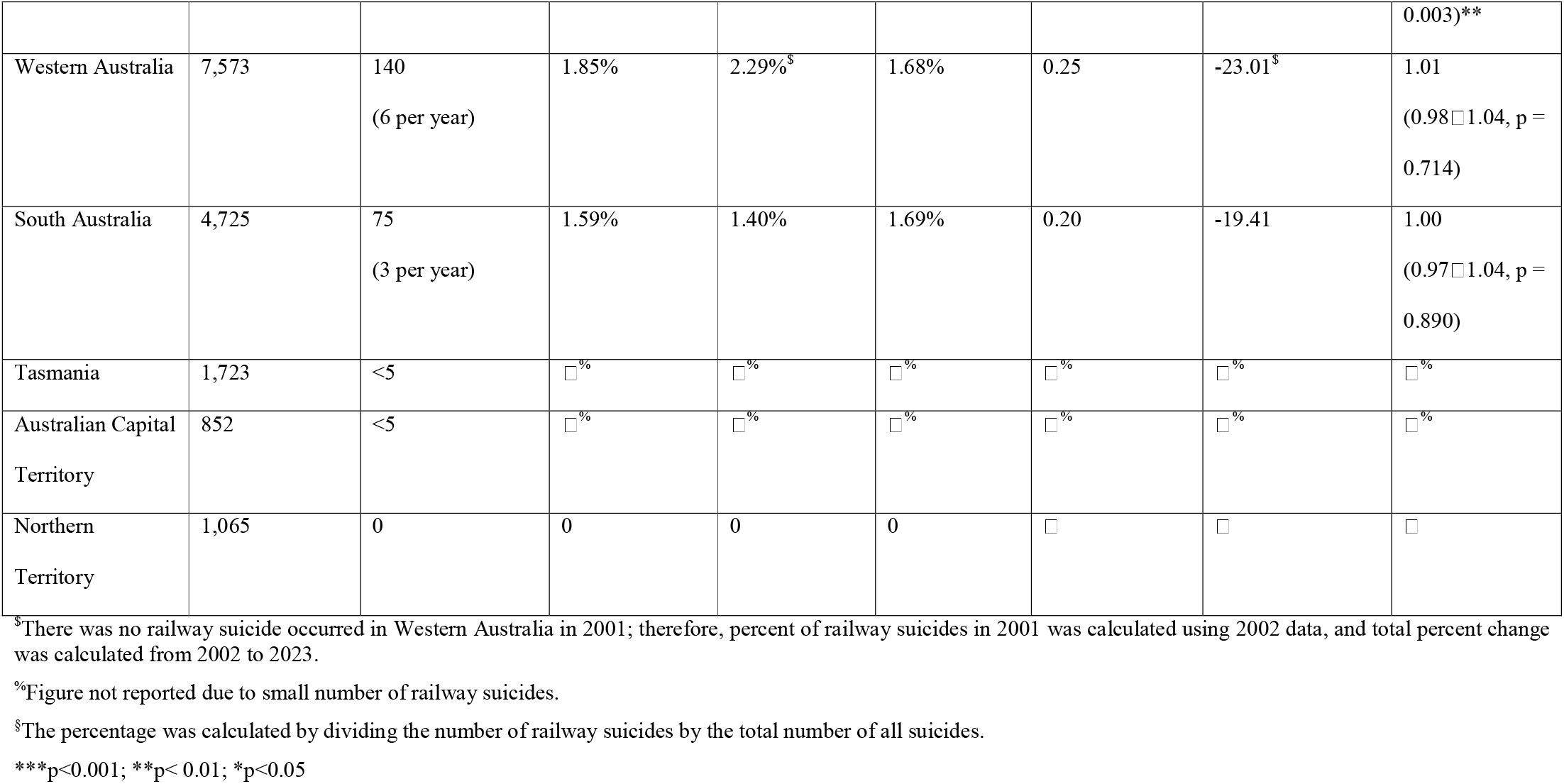
Railway suicide rates and trends across Australian states and territories (2001-2023)

### Change points for railway suicide rates

One change point was identified in Australian railway suicide rates (Table 2). Between 2001 and 2017, the rates decreased at a non-significant level. Between 2017 and 2023, the rates decreased significantly by 8.8% per year (95% CI −24.5 □-3.3) from 0.34 to 0.20 per 100,000 persons (−39.9% over this period).

**Table 2.** Change points for railway suicide in Australia and across states (2001-2023)

| Period | Annual percentage change | 95% confidence interval | p value | Total percent change in rates for each period |
| --- | --- | --- | --- | --- |
| **Australia (1 change point)** |  |  |  |  |
| 2001–2017 | -0.91 | -2.14 □ 7.25 | 0.387 | -23.52 |
| 2017–2023 | -8.79 | -24.54 □ -3.29 | <0.001 | -39.86 |
| **Victoria (1 change point)** |  |  |  |  |
| 2001–2018 | -1.00 | -2.51 □ 4.58 | 0.482 | -27.92 |
| 2018–2023 | -13.57 | -38.47 □ -4.30 | < 0.001 | -46.88 |
| **New South Wales (no change point)** |  |  |  |  |
| 2001–2023 | -2.79 | -4.61 □ -1.09 | 0.004 | -66.69 |
| **Queensland (no change point)** |  |  |  |  |
| 2001–2023 | -3.47 | -6.44 □ -0.73 | 0.020 | -25.19 |
| **Western Australia<sup>s</sup> (1 change point)** |  |  |  |  |
| 2002–2010 | 10.25 | 1.54 – 58.81 | 0.017 | 67.19 |
| 2010–2023 | -5.35 | -19.82 – -1.82 | 0.004 | -53.95 |
| **South Australia (no change point)** |  |  |  |  |
| 2001–2023 | 0.22 | -1.76 – 2.43 | 0.768 | -19.41 |

<sup>\$</sup>There was no railway suicide occurred in Western Australia in 2001; therefore, joinpoint analysis was conducted from 2002 to 2023.

In Victoria, there was no evident decline in railway suicide rates between 2001 and 2018 and the rates declined significantly by 13.6% per year (95% CI −38.5 □-4.3) from 0.60 to 0.32 per 100,000 persons (−46.9%) between 2018 and 2023. In Western Australia, railway suicide rates increased by 10.3% per year (95% CI 1.5 □58.8) from 0.31 to 0.52 per 100,000 persons (67.2%) between 2002 and 2010, and decreased by 5.4% per year (95% CI −19.8 □-1.8) from 0.52 to 0.24 per 100,000 persons (−54%) between 2010 and 2023. No change points were identified for New South Wales, Queensland, and South Australia.

## Discussion

This study used 23 years of coronial data to examine railway suicide trends and change points in Australia and across states/territories. We found that railway suicide rates decreased by 54% in Australia between 2001 and 2023 (an average −2% per year). Substantial decreases were observed in Victoria (−62%, −3% per year), New South Wales (−67%, −3% per year), and Queensland (−25%, −3% per year). Nationally, one significant change point was identified: between 2017 and 2023 (−8.8% per year). Evident change points were also identified in Victoria (between 2018 and 2023: −13.6% per year) and Western Australia (between 2002 and 2010: 10.3% per year, and between 2010 and 2023: −5.4% per year).

The reduction of railway suicides in Australia appears to be predominantly explained by the reductions in Victoria and New South Wales (the states with higher railway suicide rates), and minimally by the reduction in Queensland. Several interventions were implemented in these states, particularly after the ministerial/stakeholder roundtables on “Suicide Safer Railways” in 2017/2018. These includes the installation of fencing at high-risk locations on the railway networks in Victoria and New South Wales (NSW Health, 2021) as well as the implementation of the “Pause.Call.Be Heard” campaign which encouraged help-seeking at railway stations across the country.

In Victoria, railway locations with suicide clusters (which accounted for 35% of all railway suicides) (Too et al., 2017b) were fenced between 2017 and 2020 (based on the fencing data from the Metro Trains Melbourne). This intervention is likely to have contributed to the reduction of railway suicides in Victoria, given the strong evidence of the effectiveness of restricting access to means in preventing suicide at high-risk locations (Pirkis et al., 2015; Too et al., 2025). Notably, approximately 60% of the Victorian railway network has been fenced by 2024 (~40% meet the standard) (Victorian State Government, 2024), compared to approximately 10% in 2016 (Too et al., 2017). In 2016, a level crossing removal project was initiated in Victoria aiming to remove 110 level crossings by 2030 (Victorian State Government, 2026). By the end of 2023, 75 level crossings had been removed, which is also likely to have contributed to the reduction in Victorian railway suicides. Consistent with this, a recent study showed that railway suicides decreased within 500m/1000m of the sites where level crossings were removed (Clapperton et al., 2022).

In New South Wales, bystander intervention at railway stations may have played a role in reducing railway suicides (Ngo et al., 2022), given most incidents occurred at stations (generally without platform barriers and with Security Control Centre which coordinates communication and responses using mobile device and CCTV) (Gregor et al., 2019). This is supported by evidence showing that increased number of video surveillance systems at railway stations was associated with a reduction in railway suicide (Too et al., 2015).

The “Pause.Call.Be Heard” campaign that was implemented as one of the national preventive initiatives was not found to directly prevent railway suicide (Too et al., 2020). However, there was an increase in help-seeking intention and behaviours among rail commuters who recalled seeing the campaign messages and visuals (Too et al., 2020). This suggests that help-seeking campaigns can be a valuable component of railway suicide prevention strategies but cautions against over-reliance on their direct influence on reducing suicide. Their impact is more likely to be upstream and preventative in nature.

The downward trend in railway suicide rates observed in Australia between 2017 and 2023 could be associated with interventions introduced around or shortly after 2017. This aligns with the timing of the roundtable meetings in Victoria and New South Wales, as well as other relevant meetings in 2017, which recommended preventive interventions (e.g., installing fencing at high-risk locations and help-seeking campaigns). These interventions, along with the progressive removal of level crossings, have been implemented intermittently/continuously since 2017 and appear to have helped sustain the reduced rates over the period. In Victoria, a change point was identified between 2018 and 2023, indicating a similar but slightly later decline in railway suicide rates compared with the national pattern. This may reflect a longer lag between implementation and observable impact on railway suicide in Victoria, which has the highest rate and with a largely unfenced/highly accessible railway network. Additionally, the earlier decline at the national level may be driven partly by the declines in other states; for example, a yearly decline in railway suicide rates was found in Western Australia between 2010 and 2023, though the factors driving this change are unknown.

### Limitations

This study has some limitations. First, railway suicide may be underestimated due to classification of suicide as deaths from other causes or undetermined intent, and not all case investigations been completed at the time we accessed the data (89% cases in 2023 were closed, National Coronial Information System, 2026). Second, preventive interventions targeting suicide more generally may have been deployed in the regions with railway infrastructure and could explain the reduction in railway suicide rate; these factors were not considered in our study. Third, COVID-19 lockdowns may have restricted access to railway stations/tracks, potentially reducing railway suicide rates; however, lower rates persisted in Australia and all states after restrictions were lifted in 2022 and 2023 (see Figure 1 for railway suicide rates in Australia and across states between 2001 and 2023). Finally, although we obtained information on initiatives/interventions from the TrackSAFE Foundation (the national organisation works to reduce deaths, injuries, and near hits on the railway network, established in 2012) and through relevant online sources/articles, we may miss initiatives/interventions took place in the country and states/territories, particularly those before 2012.

## Conclusion

National railway suicide rates have declined markedly in Australia, primarily driven by reductions in railway suicides in Victoria and New South Wales. These changes appear to have occurred after the introduction of a variety of interventions. This indicates that, while challenging, railway suicide is preventable through multisectoral collaborations and the adoption of research-informed prevention strategies. We recommend the continued implementation of interventions, including fencing installation, level crossing removal, bystander intervention, and campaigns designed to increase help seeking to reduce railway suicide.

## Acknowledgements

LST was supported by a CR Roper Fellowship and held the University of Melbourne’s Research Impetus Grant. PCFL was supported by the Research Impetus Grant. JP was supported by a NHMRC Investigator Grant (No. 2026408). We would like to thank the NCIS (database source of data) and Victorian Department of Justic and Community Safety (organisational source of data) for approving and providing the data access.

## Author contributions

## Ethical considerations

This study was approved by the Health Sciences Human Ethics Committee (the University of Melbourne, application number: 2025-23756-72991-8) and the Victorian Justice Human Research Ethics Committee (reference number: CF/14/2280).

## Consent to participate

Not applicable.

## Consent for publication

Not applicable.

## Data availability

The data used in this study are restricted to LST who was granted the data access by the Victorian Justice Human Research Ethics Committee.

