## Supplementary material for "Preventing railway suicide: learning from Australia’s success": Supplementary material.docx

Table S1. Timeline and railway suicide prevention initiatives/interventions in Australia

| **Date** | **Preventive measures/activities** |
| --- | --- |
| 2013‒2016 | The Victorian railway suicide prevention research program led by a research team [lead investigator: Lay San Too], initially based in Monash University and then the program was relocated to the University of Melbourne. Stakeholders involved key representatives from the Victorian Department of Transport, Planning and Local Infrastructure, Victorian Police, Coroners Court of Victoria, VicTrack, V/line, Public Transport Victoria, Metro Trains Melbourne, Transport Safety Victoria, and Australian Rail Track Corporation.  Stakeholder meetings were held between 2013 and 2016 to discuss knowledge gaps, preventive efforts to mitigate suicide risk on the railway networks, and the availability/accessibility of relevant data from the railway sectors. During this period, research findings when available were disseminated to stakeholders to inform railway suicide preventive strategies.  Key representatives noted that, although restricting access to means is a highly effective suicide preventive strategy, fencing the entire railway network is not feasible. To address this challenge, the research team recommended prioritising fencing at high-risk railway locations (i.e. locations frequently used for suicide/where suicide clusters have been identified). Our research identified four suicide clusters on the Victorian railway network. |
| October‒December 2016 | Community Stations Project, instigated by Public Transport Victoria, Metro Trains Melbourne, and TrackSAFE Foundation. Four railway stations across Melbourne participated in this project, implementing various interventions (arts and culture, music, food and coffee, and awareness-raising events) to improve emotional wellbeing and understanding of poor mental health, and ultimately prevent railway suicides at stations. |
| 2016‒2030 | Level crossing removal project in Victoria (removing 110 level crossings by 2030) to improve safety, reduce congestion, improve travel time reliability, and increase capacity to run more trains on the network. 11 level crossings were removed by the end of 2017, 75 removed by 2023, and 88 removed by April 2026. |
| February 2017 | Roundtable on Suicide Safer Railways in New South Wales, hosted by Lifeline Australia at the presence of the NSW Minister for Mental Health, and the NSW Minister for Transport and Infrastructure, and 29 senior representatives from government and community organisations. LST presented the research findings and provided recommendations for reducing railway suicide, highlighting installing physical barriers at high-risk railway locations. Roundtable report outlining a framework for future actions to reduce railway suicide was submitted to the Ministers. |
| November 2017 | LST met with Department of Health & Human Services, Victoria to disseminate the locations of suicide clusters on the Victorian railway network, and for them to prepare a submission for funding from the Commonwealth Australia for ‘hotspot’ suicide prevention infrastructure projects. |
| December 2017 | Roundtable on Suicide Safer Railways in Victoria, hosted by the Victorian Minister for Public Transport, Minister for Roads and Road Safety, and Minister for Mental Health at the presence of 43 senior representatives from the government, the police, railway industry, public transport, roads, and mental health sector. LST presented the research findings and provided recommendations for reducing railway suicide, highlighting installing physical barriers at high-risk railway locations. Ministerial roundtable report outlining recommendations for preventing railway suicide was submitted to the Ministers. |
| December 2017 | LST met with Head of Strategy & Project Integration, Metro Trains Melbourne to provide recommendations for them to develop strategies for reducing railway suicides. |
| November 2018 | Roundtable on Suicide Safer Railways in Queensland, hosted by the TrackSAFE Foundation and Lifeline Australia at the presence of Queensland Minister for Transport, the Queensland Mental Health Commissioner, and senior representatives from the rail industry, roads, police, and mental health sectors. |
| December 2017 to November 2018 | Pause.Call.Be Heard campaign, developed by the TrackSAFE Foundation and the Lifeline Research Foundation, run at several railway stations in Victoria. The campaign contained posters and digital billboards showing the Lifeline crisis helpline number and the “Pause.Call.Be Heard” messages. The digital billboards also contained a guided breathing exercise. |
| 2017‒2020 | Fencing was installed at 39 locations on the Victorian railway network, including all the identified high-risk railway locations (fencing information obtained from Metro Trains Melbourne). |
| 2020 | Geotargeted Pause.Call.Be Heard campaign run at prioritized areas of the railway network in Victoria. |
| 2021 | New additional corridor fencing was installed across 2.3km of NSW railway network. |
| 2022 | Geotargeted Pause.Call.Be Heard campaign run at prioritized areas of the railway network in Victoria. |
| May to June 2022 | Geotargeted Pause.Call.Be Heard campaign run at prioritized areas of the railway networks in Sydney, Adelaide, Brisbane, and Perth. |
| 2023 | Geotargeted Pause.Call.Be Heard campaign run at prioritized areas of the railway network in Victoria. |
| September 2023 | Launch of TrackSAFE’s Suicide Awareness Training (bystander intervention). This training aimed to increase rail workers’ knowledge about appropriate ways to intervene with people at risk of suicide and increase rail workers’ self-reported willingness and confidence to intervene. |
| 2024 | 650 Lifeline help-seeking signs installed at railway stations in Victoria  Geotargeted Pause.Call.Be Heard campaign run at prioritized areas of the railway network in Victoria.  Geotargeted Pause.Call.Be Heard campaign and cross track outdoor billboards installed at two railway stations in New South Wales. |
| June 2024 | Approximately 60% of the Victorian railway network has been fenced by this time (~40% meet the standards). |
| 2025 | Geotargeted Pause.Call.Be Heard campaign and Lifeline text advertising run at prioritized areas of the railway network in Victoria. |
| 2026 | Geotargeted Pause.Call.Be Heard campaign and Lifeline text advertising run at prioritized areas of the railway network in Victoria. |

Note: The TrackSAFE Foundation provided information about their railway suicide prevention activities.
